# Sleep quality does not mediate the association between chronodisruption and metabolic syndrome

**DOI:** 10.1101/2020.10.30.20223164

**Authors:** Tannia Valeria Carpio-Arias, Diana Carolina Mogrovejo Arias, Tomas Marcelo Nicolalde-Cifuentes, Estephany Carolina Tapia-Veloz, Chris I. De Zeeuw, María Fernanda Vinueza-Veloz

**Affiliations:** Research group GIANH, Escuela Superior Politécnica de Chimborazo, Riobamba, Ecuador; Public Health Research Group, Universidad de Alicante, Alicante, Spain; Faculty of Mathematics and Natural Sciences, University of Potsdam, Potsdam, Germany; School of Medicine, Escuela Superior Politécnica de Chimborazo, Riobamba, Ecuador; Department of Pharmacy and Pharmaceutical Technology and Parasitology, University of Valencia, Valencia, Spain; Department of Neuroscience, Erasmus University Rotterdam, Rotterdam, The Netherlands; Research group PSICOMED, Escuela Superior Politécnica de Chimborazo, Riobamba, Ecuador

**Keywords:** shift work, metabolic syndrome, sleep quality, chronodisruption

## Abstract

**Aims:** The objective of the present work was to determine to what extent sleep quality may mediate the association between CD, metabolic syndrome (MS) and body composition (BC).

**Methodology:** Cross-sectional study which included 300 adult health workers, 150 of whom were night shift workers and thereby exposed to CD. Diagnosis of MS was made based on Adult Treatment Panel III criteria. Sleep quality was measured using the Pittsburgh Sleep Quality Index. Body mass index (BMI), fat mass percentage, and visceral fat percentage were measured as indicators of BC. Data were analyzed using logistic, linear regression and structural equation models.

**Results:** The odds of health workers exposed to CD to suffer from MS was 22.13 (IC_95_ 8.68 - 66.07) when the model was adjusted for age, gender, physical activity and energy consumption. CD was also significantly associated with an increase in fat mass and visceral fat percentages, but not to BMI. Surprisingly, there was not enough evidence supporting the hypothesis that sleep quality contributes to the association between CD and MS or BC.

**Conclusions:** Sleep quality does not mediate the negative effects of CD on health and BC.

## 1. Introduction

Chronodisruption (CD), which concerns a significant disturbance of the circadian organization of physiology, metabolism and behavior, can on the one hand be caused by factors related to diet and activity (1), but in turn also lead to disturbances in metabolic activity (2), potentially generating a negative vicious circle. Indeed, lack of synchronization of internal biological systems relative to external, i.e., environmental changes may have general negative effects on health and eventually result in chronic diseases such as metabolic syndrome (MS) (2-7). MS is characterized by co-occurrence of interrelated metabolic factors such as insulin resistance (diabetes type II), abdominal obesity and atherogenic dyslipidemia (8,9). As sleep has been recognized as one of the modulators of metabolic homeostasis, sleep disorders may form one of the risk factors contributing to MS (10, 11).

CD can be artificially induced by working in night shifts, which involves non physiological rest schedules and exposure to artificial light at night (3). In line with the association between CD and metabolic symptoms (2-7), night shift workers have an increased risk of suffering from not only MS, but also from cardiovascular diseases including coronary events, myocardial infarction and ischemic stroke (12-14). Likewise, night shift workers, who also show increased odds of smoking and consuming higher amounts of caffeine (2,6,7), suffer from body weight increase (15), often are faced with severe sleep disturbances (10,11). These findings raise the possibility that hampered quality of sleep is one of the main factors contributing to the link between CD and MS. To investigate this hypothesis we set out to analyze the associations between sleep quality, BC and MS in a group of night shift workers and a group of regular day workers for control.

## 2. Subjects, materials and methods

### 2.1 Subjects

A non-experimental cross-sectional design was implemented. A total of 300 volunteers who worked in health care centers in the central area of Ecuador was recruited. The sample comprised 150 night shift workers and 150 day workers, including adult men and women between 18 and 60 years old. A subject was considered a night shift worker if he or she was working 8 hours or more during the night, from 7:00 pm to 06:00 am (when the sun remains hidden in Ecuador), over the last six months. All data were collected by trained and qualified personnel.

### 2.2 Ethical considerations

The study was performed in accordance with the ethical guides of the Helsinki Declaration of Human Studies. The written informed consent was obtained from all the participants of the study. Patient data were codified to guarantee anonymity. The study was approved by a institutional review committee of the Research Institute of the Escuela Superior Politécnica de Chimborazo.

### 2.3 Measurements

#### 2.3.1 Anthropometric and body composition (BC) measurements

Height was measured with a rigid and in-extensible wall tape of 60 to 210 cm at an accuracy resolution of 0.1 cm; weight was measured with a 50 gr precision scale; and waist circumference was measured with a 0.6 mm wide metal tape taking the midpoint of the distance between the edge of the iliac crest and the ribcage. Subjects were measured with the least amount of clothing possible, 2 to 6 hours post prandial. Body mass index (BMI) was obtained using the equation: BMI = weight (kg) / height (m)^2^. Fat mass and visceral fat were measured with a scale of electrical bioimpedance, brand SECA, with a reading range of 0 to 120 kg, and a precision of 100 grams.

#### 2.3.2 Sleep quality

The Spanish version of the Pittsburgh Sleep Quality Index (PSQI) was used to determine sleep quality (16). This index consists of 19 self-evaluated items, which are grouped into seven items, including subjective sleep quality, sleep latency, sleep duration, habitual sleep efficiency, presence of a sleep disorder, use of hypnotic medications, and daytime dysfunction. Each of these items receives a score of 0-3 points. The total score is the result of the sum of the 7 components, so the test can give a total score of 21 points.

#### 2.3.3 Caloric intake

Caloric intake was calculated based on the amount of food intake. To measure food intake participants filled out a food diary for every day of the last week. Data were analyzed using the Ecuadorian food composition table, obtaining average amounts of caloric intake measured in kilo calories (kcal) for each day for each subject. Caloric intake was then calculated averaging kcal consumed during the week. Prior registration, participants were trained by a nutritionist on how to fill out the diary. Participants were also provided with a telephone number for communication in case of doubts.

#### 2.3.4 Biochemical parameters

Triglycerides, HDL cholesterol and fasting glycaemia values were obtained from clinical records. These values were used only if measurements were made no more than 12 weeks before the other measurements were made.

#### 2.3.5 Systolic and diastolic blood pressure

A SECA sphygmomanometer was used to measure blood pressure (BP). Systolic blood pressure (SBP) and diastolic blood pressure (DBP) were measured in mm Hg following international recommendations for conventional, ambulatory and home blood pressure measurements (17). BP was measured twice with a five-minute time lapse between each measurement. If the difference between measurements was ≥ 5 mm Hg, a third measurement was made 10 minutes after the subject kept rest.

#### 2.3.7 Metabolic syndrome (MS)

The Third Report of the Expert Panel on Detection, Evaluation, and Treatment of High Blood Cholesterol in Adults (also referred to as the Adult Treatment Panel III or ATP III criteria) was used to diagnose MS (8). MS was diagnosed when three or more of the following criteria were met: 1) abdominal obesity (waist circumference in men > 102 cm and in women > 88 cm); 2) triglycerides ≥ 50 mg/dl; 3) HDL cholesterol (in men < 40 mg/dl and in women < 50 mg/dl); 4) BP ≥ 130/85 mm Hg; and 5) fasting glucose ≥ 110 mg/dl.

#### 2.3.1 Physical activity

The short version of the International Physical Activity Questionnaire (IPAQ) for adults was used to measure the physical activity of the subjects in the study sample (18). The IPAQ provides information on the time spent walking, activities of moderate and vigorous intensity, and sedentary behavior during the 7 days prior to the survey. Accordingly, the subjects were classified into an active, moderately active, or sedentary group.

### 2.4 Statistical analyses

Results were expressed as means and standard deviations for quantitative variables with normal distribution and frequencies and percentages for categorical variables. Differences regarding age, gender, physical activity, sleep quality, energy consumption, body composition, ATP III criteria and MS were determined using Chi^2^ test, t test, and Wilcoxon signed rank test, depending on the type of the variable (see Annex 1). To determine the association between MS and night shift work we implemented binomial logistic regression models, considering MS as a response variable and night shift work as an explanatory variable. Models were adjusted for age, gender, physical activity, and energy consumption. To determine the association between BMI, fat mass, visceral fat and shift work we implemented linear regression models, considering BMI, fat mass, and visceral fat as response variables and shift work as an explanatory variable. Models were adjusted for age, gender, physical activity, and energy consumption.

In order to find out whether sleep quality contributes to the association between night shift work, MS and BC we followed the causal steps approach outlined by Baron & Kenny in 1986 (19). Mediation analysis allows the estimation of a direct effect relating the independent to the depended variable and a mediated effect (indirect) by which the independent variable indirectly affects the dependent variable through the hypothesized mediating variable. We defined MS as the dependent variable, shift work as the independent variable and sleep quality as the hypothesized mediating variable. BC was modeled as the latent factor determined by BMI, fat mass and visceral fat as indicators. To perform mediation analysis, we used structural equation modeling (SEM). Mediation analysis was performed using Lavaan v0.6-5 (20). All statistical analyses were performed using R v3.6.3 (21).

## 3. Results

### 3.1 General characteristics of the sample

The sample included 300 adult men and women (65% and 35%, respectively), half of whom were exposed to CD, because they were night shift workers (n = 150 vs. n = 150). Mean age of people of the sample was 38.11 years (SD 7.83). Distribution of gender was similar among both groups of workers (*p* = 0.09; Table 1; Supplementary tables 1). People who worked in shifts were older, more sedentary, and had a higher energy intake (kcal/day) than people who did not work in shifts (all *p* < 0.05; Table 1; Supplementary tables). Shift workers also had a higher BMI, percentage of fat mass and visceral fat than those non-shift workers (all *p* < 0.05; Table 1; Supplementary tables).

**Table 1.**
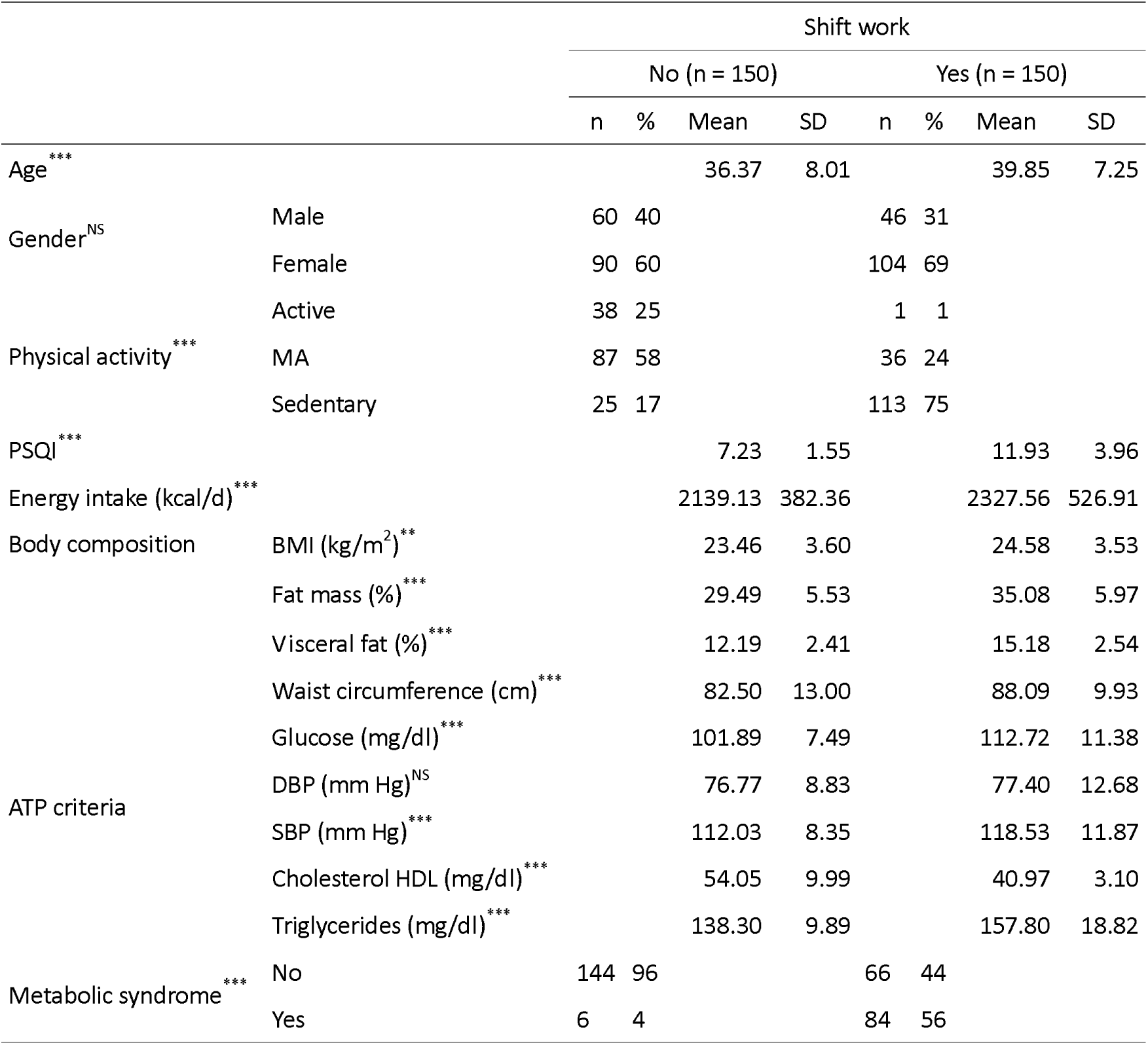
General characteristics of the sample. Sample included 300 people, 150 who worked in shifts and 150 who did not. General characteristics of the population are shown in relation to the presence or absence of shift work. *Abbreviations and nomenclature: n = number; kg = kilograms; kg/m*^*2*^ *= kilograms per squared meter; kcal/d = kilocalories per day; SD = standard deviation; BMI = body mass index; % = percentage; MA = moderately active; PSQI = Pittsburgh Sleep Quality Index; * = p < 0.05; ** = p < 0.010; *** = p < 0.001; NS = not significant*

Moreover, distribution of MS was statistically significantly different among both groups of workers (*p* < 0.001; Table 1; Supplementary Table 1). Percentage of people who met the ATP III criteria for MS was higher in the group of shift workers in comparison to not-shift workers (56% vs. 4%; Table 1). Table 1 shows the breakdown of MS criteria among workers. Except for DBP, mean values of waist circumference, glucose, SBP, and triglycerides of shift workers were statistically significantly higher than those of non-shift workers (all *p* < 0.05; Table 1; Supplementary tables). Moreover, cholesterol HDL was significantly lower (*p* < 0.001; Table 1).

### 3.2 Both metabolic syndrome and body composition are associated with shift work

Health workers that were exposed to CD showed higher odds of MS in comparison to people who were not exposed to CD (OR = 30.55; CI_95_ 13.70 – 81.64) (Table 2, non-adjusted model). This OR decreased to 22.13 (IC_95_ 8.68 – 66.07) when the model was adjusted for age, gender, physical activity and calorie intake (Table 2, adjusted model). In order to study the relationship between BC and CD we assessed the association between BMI, percentage of fat mass and visceral fat as indicators of BC and night shift work. From the three indicators, only BMI did not show a statistically significant association with shift work when the model was adjusted for age, gender, physical activity and calorie intake (Table 2, adjusted model). Working in night shifts increased percentage of fat mass by 3.32% (95% CI 1.71:4.92) and percentage of visceral fat by 2.24% (95% CI 1.54:2.96).

**Table 2.** Association between working shift and MS and body composition. Model was adjusted for age, gender, physical activity and energy consumption. Abbreviations: OR = odds ratio; IC = confidence interval; LI = lower limit; Ul = upper limit; p = p-value

| Variables | Unadjusted model |  |  |  |  | Adjusted model |  |  |  |  |
| --- | --- | --- | --- | --- | --- | --- | --- | --- | --- | --- |
| | OR | $\beta$ | 95% IC | | *p* | OR | $\beta$ | 95% IC | | *p* |
|  |  |  | LI | UI |  |  |  | LI | UI |  |
| **MS (yes)** | 30.55 |  | 13.70 | 81.64 | < 0.001 | 22.13 |  | 8.68 | 66.07 | < 0.001 |
| **BMI (kg/m<sup>2</sup>)** |  | 1.11 | 0.30 | 1.92 | 0.007 |  | 0.06 | -0.91 | 1.03 | 0.899 |
| **Fat mass (%)** |  | 5.58 | 4.27 | 6.89 | < 0.001 |  | 3.32 | 1.71 | 4.92 | < 0.001 |
| **Visceral fat (%)** |  | 2.99 | 2.42 | 3.55 | < 0.001 |  | 2.24 | 1.54 | 2.96 | < 0.001 |

### 3.3 Sleep quality does not mediate the association between shift work and metabolic syndrome

Next, we wanted to investigate whether sleep quality could mediate the effect of CD on MS. Accordingly, we tested this hypothesis using SEM (see Methods). We observed that most of the effect of shift work on MS was transmitted directly and not by mean of sleep quality. For instance, from the total standardized total effect (0.58, *p* < 0.001), all of it was transmitted directly (Table 3). Moreover, mediated effect (indirect effect) did not reach statistical significance (Table 3).

**Table 3.** Mediation analysis for MS and BC. Tested mediator was PSQI. Model was adjusted for age, gender, physical activity and energy consumption. Model was fitted using SEM. Body composition was modeled as a latent factor using BMI, fat mass, and visceral fat. Abbreviations: MS = metabolic syndrome; BC = body composition; PSQI = Pittsburgh Sleep Quality Index; p = p-value; <- = depends on

| ***Pathway*** | ***Standardized est.*** | ***p*** |
| --- | --- | --- |
| **Mediation for MS** |  |  |
| MS <- shift work (total effect) | 0.58 | < 0.001 |
| MS <- shift work (direct effect) | 0.59 | < 0.001 |
| MS <- PSQI (indirect effect) | -0.02 | 0.710 |
| **Mediation for BC** |  |  |
| BC <- shift work (total effect) | 0.36 | < 0.001 |
| BC <- shift work (direct effect) | 0.33 | < 0.001 |
| BC <- PSQI (indirect effect) | -0.03 | 0.325 |

### 3.4 Sleep quality does not mediate the association between shift work and body composition

Finally, we wanted to investigate whether sleep quality could mediate the effect of shift work on BC. Accordingly, we tested this hypothesis using SEM (see Methods). We modelled BC as a latent factor using as indicator BMI, fat mass and visceral fat as indicators. We observed that most of the effect of shift work on BC was transmitted directly and not by mean of sleep quality. For instance, from the total standardized total effect (0.36, *p* < 0.001), 0.33 of it was transmitted directly (Table 3). Moreover, mediated effect (indirect effect) did not reach statistical significance (Table 3).

## 4. Discussion

The current study on health workers subject to day-night shifts investigated the association between CD, MS, and BC, and evaluated whether sleep quality mediates their relationship. Our findings suggest that CD increases the odds of MS and changes in BC in that it increases the percentage of both fat mass and visceral fat. Interestingly, there was not sufficient evidence indicating that sleep quality mediates the deleterious impact of CD on health and BC.

### 4.1. Shift work and metabolic syndrome

We observed a significant association between CD and MS. The odds of MS of workers exposed to CD was around 22 times higher than the odds of workers who were not exposed to CD. Our findings are in agreement with those of two recent studies, a meta-analysis and a systematic review (22,23). Moreover, the results of these studies suggest that the effect of CD on MS largely depends on the time in which a person has started to be exposed to night shift work; being more deleterious if it started earlier in life. Taken together, we can conclude that night shift work should be monitored, especially from the perspective of occupational risk and care of professionals exposed to CD.

We also observed that the occurrence of MS was higher in night shift workers compared to day workers (56% versus 4%). According to the latest available data from a national survey in Ecuador more than 2 million people between 20 and 59 years of age (near 23%) suffer from MS (24). Because MS constitutes a serious public health concern, identification of vulnerable groups such as night shift workers is a necessary contribution to Ecuadorian public health that requires special attention.

### 4.2. Chronodisruption is related to less healthy parameters associated with body composition

Our study shows that people who are exposed to CD have less healthy habits. In fact, they were more sedentary and showed a higher energy intake compared to day workers. These findings are in agreement with those of two recent studies suggesting that night shift workers require interventions to improve their life-style habits (25,26). For example, nurses working in night shifts were found to have a higher energy consumption than required by their nutritional needs, causing excessive weight gain (25). Yet, some other studies indicate that the relations described above might be more subtle or at least be subject to compensatory behavior. Indeed, the study by Hulsegge and colleagues found no significant differences in the frequency of food intake or quality of food among night shift and day workers, but they did find differences in the quality of snacks a person consumed depending on whether they worked during the day or at night (27). People tended to consume healthier snacks when the work-shift occurred during daytime, compensating for the more unhealthy night shifts. These findings indicate that it is necessary to deeply understand eating habits of shift workers in order to understand the factors that control their body composition.

Night shift workers in the current study also presented less healthy values with respect to various indicators of BC, which included BMI, percentage of fat mass and visceral fat. Classically, studies on the regulation of body weight have almost exclusively focused on caloric intake and energy expenditure. However, evidence is emerging that energy regulation may be linked to circadian clock disruption and CD, suggesting that the timing of food intake itself could play an important role in weight control (28). This notion is supported by the observation that suppression of melatonin, which occurs during shift work, induces insulin resistance, glucose intolerance as well as sleep disorders, together promoting obesity (29).

Night shift work in combination with overweight and obesity often lead to an increased waist circumference, so called abdominal obesity (22). Waist circumference today is considered a good risk marker for chronic diseases such as high blood pressure, type 2 diabetes and cardiovascular disease (30). Thus, measuring waist circumference is important when assessing the nutritional status of patients who perform night shifts. In the present study we also found a significant relationship between CD and body fat mass as well as visceral fat percentage. We found that the percentages of body fat mass and visceral fat are significantly higher in night shift workers. These findings are similar to those of the work of Son 2015 (31) and Sugiura 2020 (32), who also found an association between these parameters and inadequate physical activity as well as an increased risk of chronic diseases such as atherosclerosis.

### 4.3. Sleep quality does not mediate the association with metabolic syndrome or body composition in shift workers

Surprisingly, our findings suggest that sleep quality does not contribute to the association between CD and MS or BC in night shift workers. Our results contrast previous studies in the general population that have suggested that without taking night shift work into consideration, sleep habits are associated with overweight and obesity (29). These contradictions may be explained by differences in the duration of exposure to night shift work (33,11,34), unbalance of gender distribution among the comparison groups, or differences related to dietary habits, such as breakfast frequency 2019 (35). Even though alterations of the metabolism can result from alterations in the sleep habits, napping can act as a protective factor for both weight gain and arterial hypertension (36,37). Thus, it will be interesting to study the impact of napping on other chronic conditions such as MS.

### 4.4. Strengths, limitations and methodological considerations

Given that the current study is the first of its kind in Ecuador, it has the potential to inspire further studies of MS and associated risk factors in the surrounding South American territories. Future studies should take into account various limitations we could not overcome. For example, we defined people exposed to CD as people who work in night shifts for at least 8 hours in the last six months (see Methods). We did not measure the number of consecutive night shifts, exact duration of shifts or napping habits of night shift workers, which could have an impact on our conclusions (38,36,37). Moreover, a more objective way to measure sleep quality could also be implemented. Thus, there are several improvements that could be implemented in future studies to control for potential caveats of the present study.

### 4.5. General implications and recommendations

More studies should be performed to explain the current findings in a more comprehensive manner. Further research should focus on investigating the influence of genetic, physiological, environmental as well as life-style factors on shift work, MS, BC and effects of sleep habits. Together with a more objective way to measure sleep quality could also be implemented. The findings of this study may yield important implications for public health policies. For instance, people working in night shifts may want to consider to adjust their life style and/or to change their job after having lived in a reversed circadian rhythm for a longer period of time.

## 5. Conclusions

Our findings suggest that CD increases the odds of MS and changes in BC in that it increases the percentage of both fat mass and visceral fat. Interestingly, there was not sufficient evidence indicating that sleep quality mediates the deleterious impact of CD on health and BC.

## Data Availability

Data that support the findings of this study are available on request. Data are not publicly available due to privacy or ethical restrictions based on Ecuadorian general data protection regulations.

## Acknowledgments

The authors are grateful for the collaboration of the volunteers who participated in this project. We are also grateful to M.Sc. María Fernanda Zerón and Ph.D. María Izquierdo for their comments and M.Sc. Paulina Orozco MsC for her technical support. In the same way, support of host institutions of each of the authors is appreciated. CIDZ is funded by the Dutch Organization for Medical Sciences (ZonMw), Life Sciences (ALW-ENW-Klein), European Research Council (ERC-adv and ERC-PoC), Medical Neuro-Delta, and LSH-NWO (Crossover, INTENSE).

## Limitation of liability

The authors declare that all points of view expressed in this work are their entire responsibility and not the institution in which they work, or the source of funding.

## Sources of financing

The execution of this project was not funded by any external funds. CIDZ is funded by The Dutch Organization for Medical Sciences, Life Sciences, as well as Social and Behavioral Sciences, the ERC-adv, ERC-POC and LISTEN programs of the EU, Medical NeuroDelta Program, and the NIN-Vriendenfonds voor Albinism.

## References

1. Garaulet MOJMJ. The chronobiology, etiology and pathophysiology of obesity. Physiol Behav 2010;34(12):1667–83.

2. Erren TC, Reiter RJ. Defining chronodisruption. J Pineal Res 2009;46(3):245–7.

3. Ramin C, Devore EE, Wang W, Pierre-Paul J, Wegrzyn LR, Schernhammer ES. Night shift work at specific age ranges and chronic disease risk factors. Occup Environ Med 2015;72(2):100–7.

4. Ferri P, Guadi M, Marcheselli L, Balduzzi S, Magnani D, Di Lorenzo R. The impact of shift work on the psychological and physical health of nurses in a general hospital: A comparison between rotating night shifts and day shifts. Risk Manag Healthc Policy 2016;9:203–11.

5. Proper KI, Van De Langenberg D, Rodenburg W, Vermeulen RCH, van der Beek AJ, van Steeg H, et al. The relationship between shift work and metabolic risk factors: A systematic review of longitudinal studies. Am J Prev Med 2016;50(5):e147–57.

6. Canuto R, Garcez AS, Olinto MT. Metabolic syndrome and shift work: a systematic review. Sleep Med Rev 2013;17(6):425–431.

7. Wang F, Zhang L, Zhang Y, Zhang B, He Y, Xie S, et al. Meta-analysis on night shift work and risk of metabolic syndrome. Obes Rev 2014;15(9):709–20.

8. Huang PL. A comprehensive definition for metabolic syndrome. DMM Dis Model Mech 2009;2(5-6):231–7.

9. Expert Panel on Detection, Evaluation, and Treatment of High Blood Cholesterol in Adults. Executive Summary of The Third Report of The National Cholesterol Education Program (NCEP) Expert Panel on Detection, Evaluation, And Treatment of High Blood Cholesterol In Adults (Adult Treatment Panel III). JAMA 2001;285(19):2486–2497.

10. Wickwire EM, Geiger-Brown J, Scharf SM, Drake CL. Shift Work and Shift Work Sleep Disorder: Clinical and Organizational Perspectives. Chest 2017;151(5):1156–1172.

11. Koren D, Dumin M, Gozal D. Role of sleep quality in the metabolic syndrome. Diabetes Metab Syndr Obes 2016;9:281–310.

12. Vyas MV, Garg AX, Iansavichus AV, Costella J, Donner A, Laugsand LE, et al. Shift work and vascular events: systematic review and meta-analysis. BMJ 2012;345:e4800.

13. De Bacquer D, Van Risseghem M, Clays E, Kittel F, De Backer G, Braeckman L. Rotating shift work and the metabolic syndrome: a prospective study. Int J Epidemiol 2009;38:848–54.

14. Pietroiusti A, Neri A, Somma G, Coppeta L, Iavicoli I, Bergamaschi A, et al. Incidence of metabolic syndrome among night-shift healthcare workers. Occup Environ Med 2010;67:54–7.

15. van Drongelen A, Boot CR, Merkus SL, Smid T, van der Beek AJ. The effects of shift work on body weight change - A systematic review of longitudinal studies. Scand J Work Environ Heal 2011;37(4):263–75.

16. Macías JA, Royuela A. La versión española del Índice de Calidad de Sueño de Pittsburgh. Inf Psiquiatr 1996;146:465–72.

17. O’Brien E, Asmar R, Beilin L, Imai Y, Mallion JM, Mancia G, et al. European Society of Hypertension recommendations for conventional, ambulatory and home blood pressure measurement. J Hypertens. 2003;21(5):821–848.

18. Mantilla Toloza SC, Gómez-Conesa A. El Cuestionario Internacional de Actividad Física. Un instrumento adecuado en el seguimiento de la actividad física poblacional. Rev Iberoam Fisioter y Kinesiol 2007;10(1):48–52.

19. Baron Reuben, Kenny D. The Moderator-Mediator Variable Distinction in Social Psychological Research: Conceptual, Strategic, and Statistical Considerations. J Pers Soc Psychol 1986;51(6):1173–82.

20. Rosseel, Y. Lavaan: an R package for structural equation modeling. J. Stat. Softw 2012;48(2):1–36

21. R Core Team. R: A language and environment for statistical computing. R Foundation for Statistical Computing, Vienna, Austria 2019. URL https://www.R-project.org/.

22. Sun M, Feng W, Wang F, Li P, Li Z, Li M, et al. Meta-analysis on shift work and risks of specific obesity types. Obes Rev 2018;19(1):28–40.

23. Saulle R, Bernardi M, Chiarini M, Backhaus I, La Torre G. Shift work, overweight and obesity in health professionals: a systematic review and meta-analysis. Clin Ter 2018;169(4):189–97.

24. Freire W, Ramírez M, Belmont P, Mendieta MJ, Silva KM, Romero N, et al. Resumen Ejecutivo. Tomo I. Encuesta Nacional de Salud y Nutrición del Ecuador. ENSANUT-ECU 2011-2013 Ministerio de Salud Pública/Instituto Nacional de Estadística y Censos. Quito, Ecuador 2013.

25. Peplonska B, Kaluzny P, Trafalska E. Rotating night shift work and nutrition of nurses and midwives. Chronobiol Int 2019;36(7):945–954.

26. Flahr H, Brown WJ, Kolbe-Alexander TL. A systematic review of physical activity-based interventions in shift workers. Prev Med Rep 2018;10:323–331.

27. Hulsegge G, Boer JM, van der Beek AJ, Verschuren WM, Sluijs I, Vermeulen R, et al. Shift workers have a similar diet quality but higher energy intake than day workers. Scand J Work Environ Health 2016;42(6):459–468

28. Arble DM, Bass J, Laposky AD, Vitaterna MH, Turek FW. Circadian timing of food intake contributes to weight gain. Obesity 2009;17(11):2100–2.

29. Cipolla-Neto J, Amaral FG, Afeche SC, Tan DX, Reiter RJ. Melatonin, energy metabolism, and obesity: A review. J Pineal Res 2014;56(4):371–81.

30. Chico-Barba G, Jiménez-Limas K, Sánchez-Jiménez B, Sámano R, Rodríguez-Ventura AL, Castillo-Pérez R, et al. Burnout and metabolic syndrome in female nurses: An observational study. Int J Environ Res Public Health 2019;16(11).

31. Son M, Ye BJ, Kim JI, Kang S, Jung KY. Association between shift work and obesity according to body fat percentage in Korean wage workers: data from the fourth and the fifth Korea National Health and Nutrition Examination Survey (KNHANES 2008-2011). Ann Occup Environ Med 2015;27:32.

32. Sugiura T, Dohi Y, Takagi Y, Yoshikane N, Ito M, Suzuki K, et al. Impacts of lifestyle behavior and shift work on visceral fat accumulation and the presence of atherosclerosis in middle-aged male workers. Hypertens Res 2020;43(3):235–45

33. Souza BB, Monteze NM, De Oliveira FLP, de Oliveira JM, de Freitas Nascimento S, Marques do Nascimento Neto R, et al. Lifetime shift work exposure: Association with anthropometry, body composition, blood pressure, glucose and heart rate variability. Occup Environ Med 2015;72(3):208–15.

34. Yu J. Relationship Between Long Working Hours and Metabolic Syndrome Among Korean Workers. Asian Nurs Res (Korean Soc Nurs Sci) 2017;11(1):36–41.

35. Kim KY, Yun JM. Analysis of the association between health-related and work-related factors among workers and metabolic syndrome using data from the Korean national health and nutrition examination survey (2016). Nutr Res Pract 2019;13(5):444–51.

36. Silva-Costa A, Griep RH, Rotenberg L. Night work and BMI: is it related to on-shift napping? Rev Saude Publica 2017;51:97.

37. Rotenberg L, Silva-Costa A, Vasconcellos-Silva PR, Griep RH. Work schedule and self-reported hypertension – the potential beneficial role of on-shift naps for night workers. Chronobiol Int 2016;33(6):697–705.

38. Harrington JM. Health effects of shift work and extended hours of work. Occup. Environ Med 2001;58:68–72.

